# Human Gene and Microbial Analyses Suggest Immunotherapy-like Mechanisms in Complete Response to Radiotherapy in Rectal Cancer

**DOI:** 10.1101/2022.07.07.22277387

**Authors:** A.K. Sulit, K. Wilson, J. Pearson, O.K. Silander, M. Michael, R. Ramsay, A. Heriot, F. Frizelle, R. Purcell

## Abstract

The treatment of rectal cancer usually involves total mesorectal excision, with preoperative chemoradiotherapy (CRT) that is aimed at downstaging tumors before surgical procedure. CRT response varies, with some patients completely responding to CRT and negating the need for surgery, and some patients not responding to the treatment at all. Identifying biomarkers of response to CRT would be beneficial in identifying whether a treatment would confer benefits to a patient while allowing them to avoid unnecessary morbidities and mortalities. While previous studies have attempted to identify such biomarkers, none have reached clinical utility which may be due to heterogeneity of the cancer. In this paper, we explored potential human gene and microbial biomarkers, and their possible contributions to the mechanisms of complete response to chemoradiotherapy. We carried out RNA sequencing on pairs of tumor and normal tissue from patients pre-surgery and analysed host gene expression and microbiome content. We discovered that the majority of enriched human genes in tumors of complete responders involve immunoglobulins, and enriched gene sets include complement and B-cell activation, and host response against viruses. This indicates involvement of immune responses in complete response to CRT. Among the enriched gene sets is the term defense response to bacterium, indicating a role of the microbiome in response to CRT. We discovered bacteria such as *Ruminococcaceae bacterium* and *Bacteroides thetaiotaomicron* to be abundant in tumors of complete responders. Both microbes have a history of being beneficial in treatment of cancers with immunotherapy, further emphasizing the role of immune responses in beneficial response to CRT. These results identify potential genetic and microbial biomarkers to CRT in rectal cancer, as well as offer a potential mechanism of complete response to CRT that may benefit further testing in the laboratory.

## Background

The incidence of young-onset rectal cancer (< 50 years) has been steadily increasing over the past two decades in some Western countries (Gandhi et al., 2017; Meyer et al., 2010; Siegel et al., 2019). The treatment of rectal cancer is based predominantly on preoperative staging, with upfront surgical resection reserved for early-stage tumors (T1-T2). Tumors that are locally advanced or have concerning features on preoperative MRI often require neoadjuvant chemoradiotherapy (CRT). The aim of neoadjuvant CRT is to downstage rectal tumors and reduce rates of local recurrence (Dayde et al., 2017; Feeney et al., 2019). The response to long-course neoadjuvant chemoradiotherapy varies; up to 20% of patients achieve a pathological complete response (pCR), up to 60% demonstrate partial response, and the remainder displaying resistance to nCRT (Dayde et al., 2017; Fischer et al., 2021b). These response rates could further be increased by total neoadjuvant therapy (Fischer et al., 2021b).

A pCR is defined as the absence of residual viable tumor cells in the resected specimen and the reliance on pathological confirmation of a complete response fails to identify patients that may benefit from organ preservation and be spared major surgery. Habr-Gama pioneered the adoption of a clinical complete response (cCR) as a surrogate for pCR (Habr-Gama et al., 2004). Patients who achieve a cCR may subsequently be managed using a “Watch and Wait” approach, thereby avoiding the morbidity associated with major resectional surgery. Although 25% of these patients show tumor regrowth by two years; the majority are amenable to salvage procedures (van der Valk et al., 2018).

Extensive work to identify clinical and biological markers of response to radiotherapy in rectal cancer has been carried out (Dayde et al., 2017; Fischer et al., 2021a; Garland et al., 2014; Ryan et al., 2016), however, no reliable biomarkers have been validated for clinical use. A robust biomarker that selects patients likely to achieve a pCR to neoadjuvant therapy, would allow for accurate identification of clinical responders and increase confidence in selecting patients amenable to non-operative management. Chemoradiotherapy is associated with significant localised and systemic side effects and has been demonstrated to negatively impact quality of life (Herman et al., 2013), and the identification of patients unlikely to benefit from conventional CRT would result in a reduction in CRT-associated side effects and targeting of patients for novel treatment strategies.

Due to the largely sporadic nature of colon and rectal cancer, environmental factors are likely to play a critical role in the development of the disease, and recent international data points to the importance of the microbiome in its development and progression (Ahn et al., 2013; Gao et al., 2015; Marchesi et al., 2011). Recent reports have also demonstrated that systemic effects of the gut microbiome may contribute to treatment response in other cancer types (Gopalakrishnan et al., 2018; Routy et al., 2018) and could be predictors of a favourable response to immunotherapy. In addition, gut microbiota have been shown to locally influence the treatment efficacy of irinotecan for colorectal cancer (Guthrie et al., 2017). However, although some studies on the protective effect of the microbiome on radiotherapy-induced toxicity have been carried out (Touchefeu et al., 2014; Vanhoecke et al., 2016), very little is known about whether or how the gut microbiome may regulate the tumor response to radiotherapy.

Here we present gene expression data from a unique cohort of pre-CRT rectal cancer tumors and their matched normal mucosa samples. Having matched tumors and normal samples accounts for interpersonal variation in gene expression and allows for a robust identification of tumor and microbial genes and molecular pathways associated with response to CRT in rectal cancer.

## Methods

### Patient cohort and characteristics

A total of 40 patients from two prospective cohorts of rectal cancer patients were used in this study: 20 patients from Christchurch Hospital, New Zealand, and 20 patients from the Peter MacCallum Cancer Centre, Melbourne, Australia. Biopsies of tumor tissue and adjacent, visually normal tissue (>10cm from tumor) were taken at colonoscopy, prior to treatment. Patient data, including staging, recurrence, metastases, treatment and histology was collected.

Patients who had received prior chemotherapy or radiation therapy for treatment of their rectal tumor were excluded from the study. This study was undertaken with ethical approval from the Health and Disability Ethics Committee of New Zealand (ethics approval number: 18/STH/40) and the Human Research Ethics Committees of Australia (ethics approval number: HREC 14/85). All participants provided written, informed consent.

### RNA extraction

Tumor and normal tissue biopsies were taken at colonoscopy and immediately frozen in liquid nitrogen and stored at -80°C. RNA extraction was carried out as detailed previously (Purcell et al., 2017). Briefly, RNA was extracted from < 20 mg of tissue using RNEasy Plus Mini Kit (Qiagen, Hilden, Germany), including DNAse treatment, following tissue disruption using a Retsch Mixer Mill. Purified RNA was quantified using a NanoDrop 2000c spectrophotometer (Thermo Scientific, Asheville, NC, USA), and stored at -80°C.

### RNA sequencing

RNA-sequencing was performed using an Illumina NovaSeq 6000 platform (Illumina, San Diego, CA, USA) to produce 150bp paired end reads, as previously described (Purcell et al., 2017). Ribo-Zero™ Magnetic Kit (Human & Bacteria) was used for ribosomal RNA depletion, and libraries were prepared using NEBNext^®^ Ultra™ RNA Library Prep Kit (New England BioLabs Inc.®). Approximately 50 million reads (15Gb raw data) were produced per sample on an Illumina NovaSeq 6000 instrument. Raw sequencing reads were deposited at NCBI SRA under BioProject ID PRJNA815861.

### RNA sequence data processing

Raw sequencing data was parsed through the Metafunc pipeline (Sulit et al., 2020), which performs read pre-processing, host gene mapping, and microbiome species identification. Further details of the computational pipeline may be found at https://gitlab.com/schmeierlab/workflows/metafunc, and complete analysis of this paper is available at https://gitlab.com/alsulit08/uoc_response_rectalca.

### Differential human gene expression analysis

Expression levels for each gene and sample were generated by the MetaFunc pipeline (Sulit et al., 2020), and DESeq2 (Love et al., 2014) was used to detect differentially expressed genes (DEGs). To detect DEGs that were significantly differentially expressed in the tumor relative to each participant’s normal tissue between groups of responders the model fitted by DESeq2 included covariates for response (complete or other), tissue type (tumor or normal), response x participant (index) and response x tissue. Care was taken to ensure the model matrix was of full rank, the model converged and that modelling assumptions were met. We considered genes differentially expressed if their p. adjust values are < 0.1. For details see https://gitlab.com/alsulit08/uoc_response_rectalca/-/tree/main/Human.

### Gene set enrichment analysis (GSEA)

From the results of this DESeq2 comparison, we generated a pre-ranked list of all resulting genes based on *p-values* and log_2_ fold-change, and these genes were used as input to GSEA analysis using *clusterProfiler* (Yu et al., 2012) with the *C5 Ontology Gene Sets collection (version 7)* from the molecular signatures database (MSigDB) (Liberzon et al., 2011; Subramanian et al., 2005). Specifically, we ranked the genes using the formula:

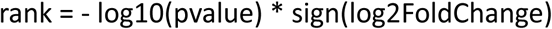

This ranking places the genes with lowest p-values and positive log_2_ fold change at the top of the list, and the genes with lowest p-values and negative log_2_ fold change at the bottom of the list. Genes at the top of the list contribute to gene sets with positive enrichment scores and genes at the bottom contribute to gene sets with negative enrichment scores. Gene sets are significantly enriched in a responder group if their p. adjust values are < 0.05.

### Differential meta-transcriptome analysis

For the microbiome dataset, we gathered raw counts of microbe taxonomies into a Phyloseq object (McMurdie and Holmes, 2013), with metadata information on their response and tumor or normal status. We then used the same model for group-specific condition effects as specified in DESeq2, to obtain differentially abundant (DA) microbes in tumor samples compared to matched normal samples, specific to complete responders compared to other responders. We performed pre-filtering of our species before the analysis, only including those within the Bacterial Kingdom, and those with at least 10 reads in 20% of our samples (**Supplementary Table 1**). Differentially abundant bacteria were species with p. adjust values < 0.1.

### Correlation between differentially expressed genes and differentially abundant bacteria in rectal cancer

Using the *rlog* transformed values for gene expression and microbial abundance obtained from DESeq2 of 87 identified DEGs and 10 DA bacterial species, we performed spearman correlation analysis between each gene and species, correcting the final p-values using Benjamini-Hochberg (BH) adjustment. We removed influential points (*species rlog > 12*) for the Spearman correlation calculation and corresponding scatter plots.

### Microbial diversity

The sample set was rarefied to 90% of the smallest sample size in the dataset and analysed in Phyloseq (McMurdie and Holmes, 2013). We obtained Observed and Shannon alpha diversities per sample and compared differences with Wilcoxon tests.

## Results

### Rectal cancer cohort

The cohort comprised 40 patients with diagnosed rectal cancer who were subsequently treated with chemo-radiotherapy (**Table 1**). One patient was subsequently excluded, due to treatment cessation for palliative care. The majority (n = 36) were treated with long course CRT (LCCRT), with either capecitabine, FOLFIRI or 5FU. Two patients did not complete LCCRT due to development of grade 3 toxicity. One patient received short-course radiotherapy, while the remaining two patients received sandwich CRT (FOLFOX). There were 12 females and 27 males, who ranged in age from 29–86 years (mean age, 62 years). Response to LCCRT was assessed histologically from surgical resection specimens, and reported using Dworak grading (Christchurch cohort) or the American Joint Committee on Cancer (AJCC) grading (Melbourne cohort). Response groups were designated as complete responders (Dworak 4/AJCC 0), near-complete responders (Dworak 3/AJCC 1), incomplete responders (Dworak 2/AJCC 2) and non-responders (Dworak 1/AJCC 3). There were five patients with complete response to LCCRT, five patients with near-complete response, 18 patients with incomplete response and eight patients who did not respond to LCCRT. In addition, three patients developed progressive disease, or died of disease, during the course of therapy, and these patients were also designated non-responders. Six patients died of the disease during a minimum follow-up period of 24 months.

**Table 1.** Characteristics of the Rectal Cancer Patient Cohort.

| **Table 1. Characteristics of the Rectal Cancer Patient Cohort** |  |  |  |  |  |
| --- | --- | --- | --- | --- | --- |
|  | **All (n=39)** | **Complete Response (n=5)** | **Near-Complete Response (n=5)** | **Incomplete Response (n=18)** | **No Response (n=11)** |
| **Age, mean ± SD** | 62.05 ± 14.55 | 66.2 ± 9.31 | 61.20 ± 4.82 | 62.44 ± 14.83 | 69.91 ± 19.29 |
| **Sex:** |  |  |  |  |  |
| **Male** | 26 (66.67%) | 4 (80%) | 4 (80%) | 13 (72.22%) | 5 (45.45%) |
| **Female** | 13 (22.22%) | 1 (20%) | 1 (20%) | 5 (27.78%) | 6 (54.55%) |
| **Died of disease:** | 5 (12.82%) | 0 (0.00%) | 0 (0.00%) | 3 (16.67%) | 2 (18.18%) |
| **Treatment:** |  |  |  |  |  |
| **LCCRT (no other data)** | 4 (10.26%) | 0 (0.00%) | 0 (0.00%) | 3 (16.67%) | 1 (9.09%) |
| **LCCRT (capecitabine)** | 26 (66.67%) | 5 (100%) | 3 (60%) | 9 (50%) | 9 (81.82%) |
| **LCCRT (5FU)** | 3 (7.69%) | 0 (0.00%) | 0 (0.00%) | 3 (16.67%) | 0 (0.00%) |
| **LCCRT (FOLFIRI)** | 2 (5.13%) | 0 (0.00%) | 0 (0.00%) | 2 (11.11%) | 0 (0.00%) |
| **Sandwich CRT** | 2 (5.13%) | 0 (0.00%) | 2 (40%) | 0 (0.00%) | 0 (0.00%) |
| **SCRT** | 1 (2.56%) | 0 (0.00%) | 0 (0.00%) | 1 (5.56%) | 0 (0.00%) |
| **Palliative** | 1 (2.56%) | 0 (0.00%) | 0 (0.00%) | 0 (0.00%) | 1 (9.09%) |
| **LCCRT: long course chemoradiotherapy** |  |  | **FOLFIRI: folinic acid, fluorouracil and irinotecan** |  |  |
| **SCRT: short course radiotherapy** |  |  |  |  |  |
| **5FU: fluorouracil** |  |  |  |  |  |

In this analysis, complete responders were compared to all other responders grouped together, and conversely, non-responders were compared to all other response groups.

### Differential gene expression and gene set enrichment between radiotherapy response groups

Differentially expressed genes (DEGs) were analysed between matched pairs of tumor and normal tissues, to account for interpersonal variation in gene expression, and then these DEGs were associated with a response group. Eighty-seven genes were found to be differentially expressed between tumor and normal samples (p. adjust < 0.1), specific to complete responders. Interestingly, the majority of these genes are associated with immunoglobulin chains, with 75 out of 87 genes having prefixes of IGH-, IGL-, or IGK (**Figure 1A**), for immunoglobulin heavy, light, and kappa, respectively. Tumor vs Normal *rlog* was plotted as transformed counts per sample of representatives from these genes and showed that, for most of these genes, complete responders cluster at high tumor-low normal values (upper left quadrant), indicating that in the complete responder group, these genes are more highly expressed in tumors compared to normal tissues (**Figure 1B and Supplementary Figure 1**). When differential gene expression analysis (DGEA) was carried out comparing non-responders to all other responders, no differentially expressed genes were identified between the two groups.

**Figure 1.**
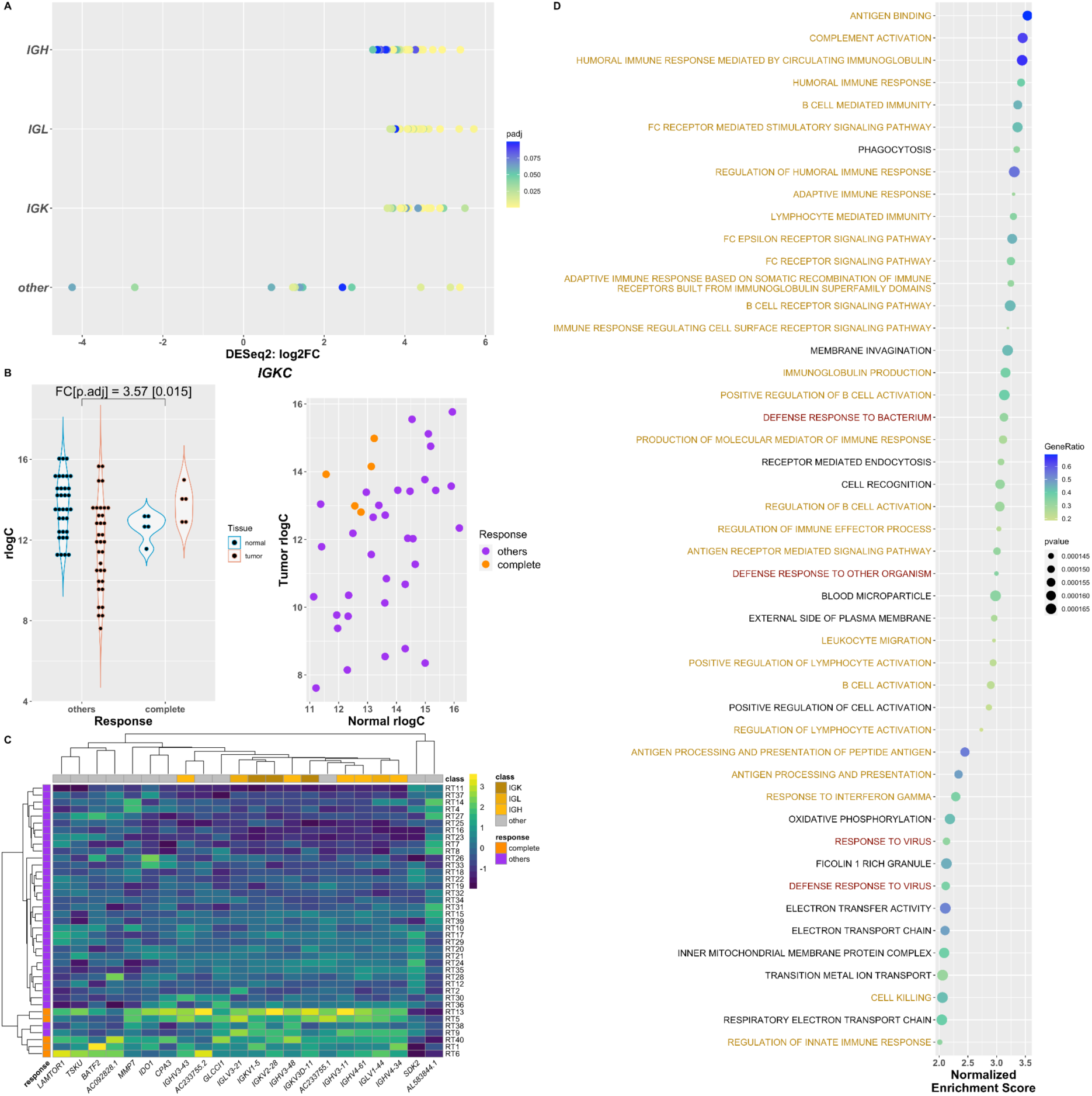
Host differential gene expression and gene set enrichment analysis. **A**. Differentially expressed genes (DEGs) classified into immunoglobulin (Ig)-related genes and other non-Ig genes. **B**. Transformed counts (rlog) of Tumor/Normal for complete responders compared to other responders of a representative DEG, *IGKC*. Left plot is Tumor vs Normal for each response category. The right plot is the tumor values and their corresponding normal values per sample by response. **C**. Heatmap of the top 10 (by p-adjust values) DEGs, which are all Ig-related genes, and the other 12 non-Ig DEGs clustered by sample and gene. **D**. Top 47 positively enriched gene sets in Tumor vs Normal of Complete Responders. All have adjusted p-values (benjamini-hochberg) of 0.0093. Gene ratio is the ratio of core enrichment genes to the gene set size.

The majority of significant differentially expressed genes were up-regulated in tumors compared to normal. The top 10 DEGs (all immunoglobulin-related), in addition to 12 non-immunoglobulin-related DEGs, can robustly distinguish complete responders from all other patients, as illustrated in **Figure 1C**.

### Host gene set enrichment analysis

Gene Set Enrichment Analysis (GSEA) was performed using genes ranked according to DESeq2 p-value and log_2_ fold-change (-/+). Gene sets relating to immune responses constitute the majority of the top 47 enriched gene sets in tumors of complete responders (**Figure 1D**). Among the gene sets with the highest enrichment scores are Complement Activation and B-Cell Mediated Immunity, consistent with the majority of the 87 differentially expressed genes in complete responders being related to immunoglobulins.

### Diversity analysis of the microbiome of rectal tumors

No significant differences were found in Observed or Shannon measures of alpha diversity and no distinct group separations by NMDS (**Supplementary Figure 2**).

### Differential abundance analysis of bacterial species between radiotherapy response groups

A similar approach was used to identify bacterial transcripts that were differentially abundant in tumor versus matched normal samples, and specific to complete responders. Analysis of differences in the tumor microbiome between response groups identified 10 bacterial species that were differentially abundant (p.adjust < 0.1) in tumor tissue compared to matched normal tissue in complete responders (**Figure 2**). Bacterial species identified included *Ruminococcaceae bacterium, Hungatella hathewayi, Bacteroides thetaiotaomicron, and Clostridium* species, which were previously reported to have a role in colorectal cancer and/or response to therapy. Plotting Tumor vs Normal *rlog* transformed counts per sample of these microbes show that there is a separation between the complete responders compared to other responders although not as pronounced as that seen in human DEGs (**Supplementary Figure 3**).

**Figure 2.**
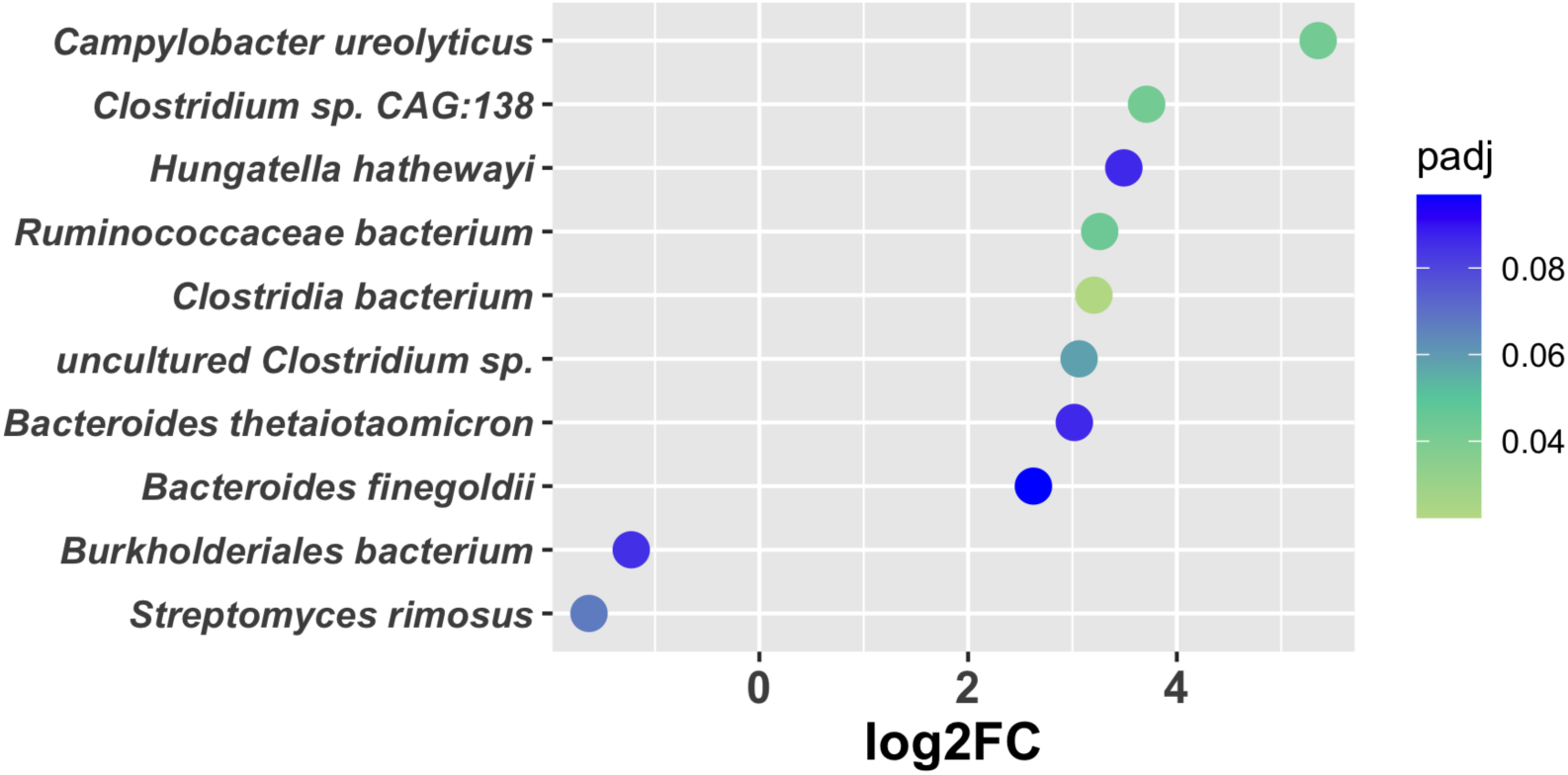
Differentially abundant bacteria in tumor samples compared to matched normal tissue, specific to complete responders. Plot of the 10 DA bacteria showing their log_2_ fold changes (x-axis) and p-adjust values by colour.

### Correlation between Host Gene Expression and Microbial Abundances

Significant correlations between differentially expressed genes and differentially abundant bacterial species were identified. Among DEGs that positively correlated with the microbial abundances, the *BATF2* gene was notable. We found positive correlations (*spearman coefficient: 0*.*355 – 0*.*549; BH adjusted p-value: 1*.*67×10*^*-4*^ *- 7*.*70×10*^*-2*^) of this gene with several of DA bacterial species, previously linked to colorectal cancer, including *Ruminococaccae bacterium*, and *Bacteroides thetaiotaomicron* (**Figure 3**).

**Figure 3.**
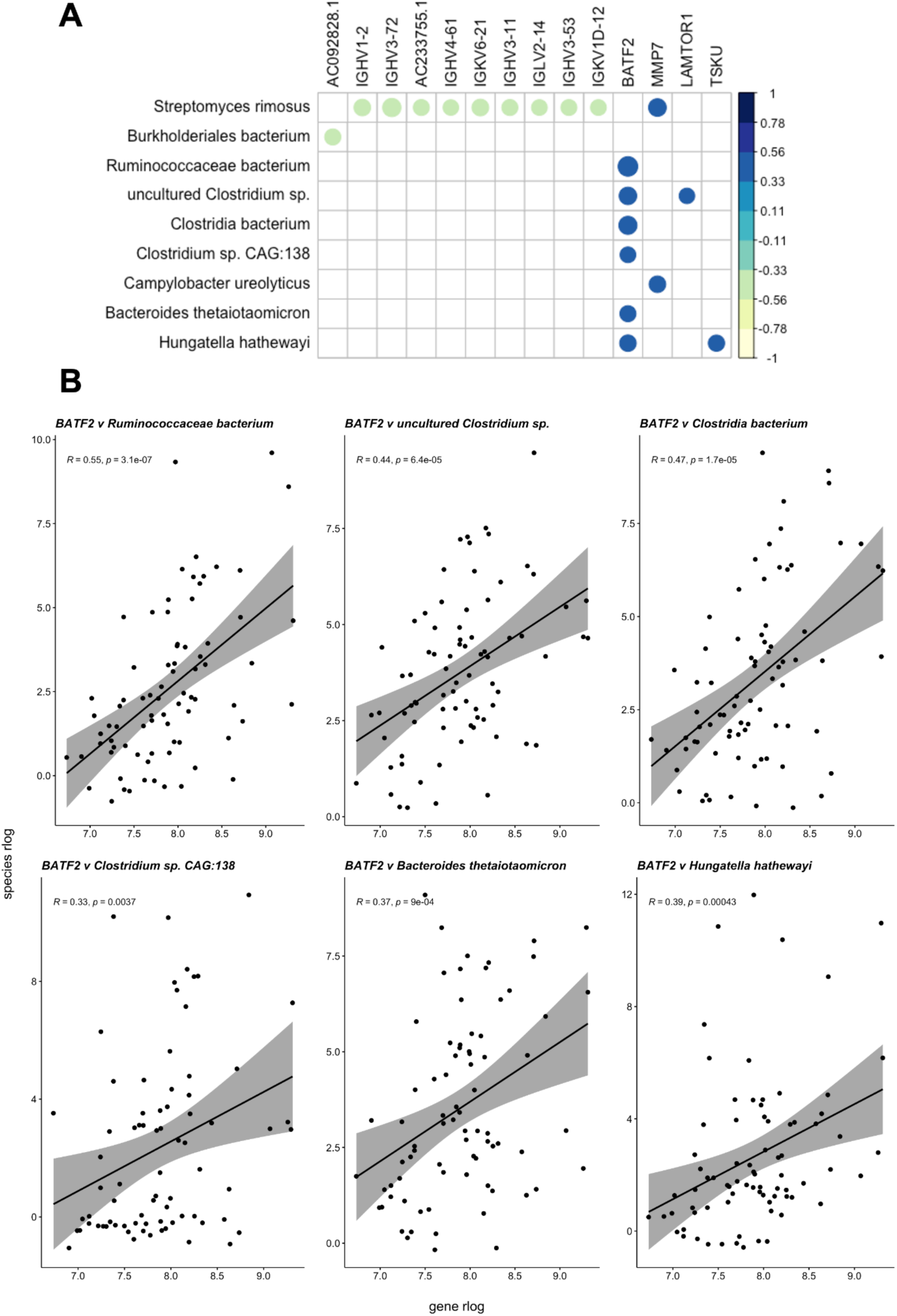
Correlations between differentially expressed genes and differentially abundant microbes. **A**. Correlation plot showing bacteria–gene correlations with at least one significant correlation (*adjusted p-value < 0*.*1*). Spearman correlation coefficients (ρ) are represented by the colour bar; blank spaces represent correlations that are not significant. **B**. *BATF2* and its positive correlations with differentially abundant bacteria. *R=spearman coefficient; p=p value calculated*.

## Discussion

The identification of predictive biomarkers of response to chemoradiotherapy will result in improved survival, a reduction of morbidities related to unnecessary treatment, and a more targeted approach to treatment for patients with rectal cancer. Previous studies have attempted to identify markers of complete response to chemoradiotherapy, and while clinicopathological and radiological features have been identified, they are limited in sensitivity and specificity (Dayde et al., 2017). Intratumoral heterogeneity contributes to lack of reproducibility between molecular biomarker studies, and as a result, no biomarker is currently in clinical use (Dayde et al., 2017; Ryan et al., 2016). In order to identify a predictive biomarker of response for rectal tumors, a mechanistic link to the underlying tumor biology is also warranted. This study identifies potential biomarkers of complete response to CRT in patients with rectal cancer, in addition to uncovering novel links between the tumor microbiome and immune response in the rectal tumor microenvironment.

Rather than a continuum of gene-expression changes from complete responders to non-responders, we observed a tight clustering of complete responders based on gene expression of mainly immune-related genes, compared to all other patients. This suggests the presence of a distinct tumor micro-environment in a group of patients that predisposes them to a complete response to radiotherapy. In our cohort, these patients showed significantly higher expression of genes responsible for complement activation and B-cell-related functions in their tumor tissue compared to adjacent normal tissue, supporting previously published reports that immunoglobulins may recognize radiotherapy-induced neo-antigens resulting in complement activation and CD8+ T-cell responses (Surace et al., 2015).

We observed enriched gene sets related to antigen presentation in our cohort of complete responders. Ionizing radiation can cause DNA mutations, which, when translated into peptides, are presented on MHC-1 molecules, eliciting a cytotoxic immune response. This has been observed in studies of other cancer types, where radiation has been shown to recruit neutrophils and monocytes, as well as promote maturation of antigen presenting cells (Krombach et al., 2019). Radiotherapy also results in mimicry of viral infections, where cytosolic DNA induces the release of type 1 interferons that recruit dendritic cells specialized in antigen presentation to CD8+ T-cells (Lhuillier et al., 2019). Our findings indicate that enhanced antigen presentation in pretreatment tumors predisposes them to successful response to CRT.

We also found a significant enrichment of the *BATF2* gene in patients who went on to have a complete response to CRT, which is consistent with findings that *BATF2* depletion in tumor compared to normal tissue is correlated with poor prognosis in colorectal cancer (Liu et al., 2015). *BATF2* is thought to be induced by type 1 interferons and play a role in viral infections (Guler et al., 2015) and may, as a result, contribute to radiotherapy-induced responses that mimic viral infection. Type-1 interferons are important players in antiviral immune responses, recruiting dendritic cells specialized in antigen presentation to CD8+ T-cells (Lhuillier et al., 2019). This is again consistent with viral infection and antigen presentation enriched gene sets we have identified in complete responders in this cohort. Furthermore, *BATF2* may also induce anti-tumor effects through the induction of CD8^+^ T-cells (Kanemaru et al., 2017).

The enrichment of immune response-related genes and gene sets in tumors of complete responders to CRT in our cohort is consistent with the findings of Sendoya et al., 2020, where they identified antigen presentation, interferon (IFN) activity, and B-cell activity to be enriched in good responders to preoperative chemoradiotherapy in locally advanced rectal cancer (LARC) patients, and that these factors may relate to good responses to the treatment via activation of an antiviral-like response and CD8+ T-cell recruitment. While their study largely used microarray gene expression analysis in their methods and used targeted sequencing of colorectal cancer (CRC) genes, this study mainly used whole RNA sequencing in our methods which also allowed us to investigate possible contributions of the microbiome.

A key finding of this study is the enrichment of gene sets relating to response to bacteria in complete responders. The role of the microbiome in therapy response has recently become a focal point for studies of different types of treatments in a variety of cancer types. Early studies of the microbiome in colorectal cancer had suggested a decrease in microbial diversity associated with tumors compared to healthy controls (Ahn et al., 2013; Gao et al., 2015; Wong et al., 2017). However, more recent studies refute this finding, where CRC samples have increased richness (Thomas et al., 2019) or are no different (Wirbel et al., 2019) compared to controls. Consistent with these more recent studies (Shi et al., 2020), we found no differences in microbiome diversity metrics between our response groups. However, we identified a number of bacterial species that were differentially abundant in tumors of complete responders, several of which had previously been implicated in CRC carcinogenesis and prognosis. *Hungatella hathewayi* has been reported to be differentially abundant in CRC (Wirbel et al., 2019), and has been reported to drive methylation of tumor suppressor genes (Xia et al., 2020) and was also found to be significantly more abundant in complete responders in our study. *Ruminococcaceae*, a putative commensal genus has been linked to the expression of T-cell recruiting chemokines (Cremonesi et al., 2018), indicating a role in tumor-killing immune activation and a low abundance has been associated with CRC (Burns et al., 2015; Yu et al., 2018). High levels of this genus may indicate a beneficial role in driving response to CRT in the tumors of complete responders through enhanced T-cell activation. Similarly, *Clostridium* species have been associated with a protective effect against CRC, due to their ability to synthesize short-chain fatty acids, notably butyrate, which can promote CRC cell apoptosis, and inhibit carcinogenesis (Zou et al., 2018).

The importance of the gut microbiome in response to immunotherapy has garnered much attention in recent years, where species of microbes have been associated with good response to immunotherapy treatments (Gopalakrishnan et al., 2018; Routy et al., 2018). Given the similarities with the known immune mechanisms at play in radiotherapy response, e.g. CD8+ T-cell activation, an overlap between the established systemic effect of the microbiome on immunotherapy efficacy and that of radiotherapy is likely. *Ruminococcaceae* and *B. thetaiotaomicron*, both enriched in complete responders in our cohort have been reported to enhance immunotherapy effects (Gopalakrishnan et al., 2018; Vétizou et al., 2015), and this mechanism may also contribute to radiotherapy-induced immune clearance of cancer cells. Indeed Gopalakrishnan et al., 2018 showed that there is a correlation between CD8+ T-cells and abundance of the *Ruminococcaceae* family, and that there are statistically more CD8+ T-cells in responders versus non-responders to immunotherapy in melanoma patients. Taken together with the associations between differentially abundant bacteria and *BATF2* expression, and the increased abundance of commensal bacteria, such as *Clostridium spp*, the activation of immune pathways in the tumor microenvironment indicates a potential role of the tumor microbiota in radiotherapy response in rectal cancer. Our findings also suggest the potential use of faecal microbiota transplants as a means of improving efficacy of CRT, in a similar manner to that reported for immunotherapy.

In this work, we identified potential predictive biomarkers and their respective biological mechanisms in the response to radiotherapy, providing a framework for future research. While this study is among the largest of its kind to investigate paired pre-treatment tumor and normal tissue from patients with rectal cancer, future studies to assess the clinical utility of incorporating immune and bacterial gene markers to stratify patients for targeted therapy are necessary in a larger prospective cohort.

## Conclusion

Our results suggest that an increase in the expression of genes contributing to immune activation in tumors compared to normal samples contributes to radiosensitivity. We hypothesize that this primes a microenvironment that activates anti-tumor responses during radiotherapy. Bacteria enriched in the tumors of complete responders in this study have previously been associated with colorectal cancer and improved efficacy of immunotherapy, and possibly contribute to the immune activation taking place in complete response to radiotherapy. These data provide future targets for biomarker validation and provide direction to investigate the mechanisms of radiotherapy response in rectal cancer.

## Data Availability

Raw sequences used in the study are available as BioProject ID PRJNA815861. The bioinformatics methodology is accessible at https://gitlab.com/schmeierlab/workflows/metafunc and further analyses at https://gitlab.com/alsulit08/uoc_response_rectalca.

https://www.ncbi.nlm.nih.gov/bioproject/?term=PRJNA815861

https://gitlab.com/schmeierlab/workflows/metafunc

https://gitlab.com/alsulit08/uoc_response_rectalca

