## Supplementary Material for "Human Gene and Microbial Analyses Suggest Immunotherapy-like Mechanisms in Complete Response to Radiotherapy in Rectal Cancer"

**Supplementary Tables**

**Supplementary Table 1. Summary statistics of sample read counts at each stage of filters applied**

| | All Species | Bacterial Species | Bacterial species (≥10 reads in ≥ 20% samples) |
|---|---|---|---|
| **Minimum** | 216192 | 196204 | 184911 |
| **Maximum** | 5931870 | 5882950 | 5845561 |
| **Mean** | 720690 | 676079 | 652713 |
| **Median** | 519818 | 481973 | 468629 |

### Supplementary Figures

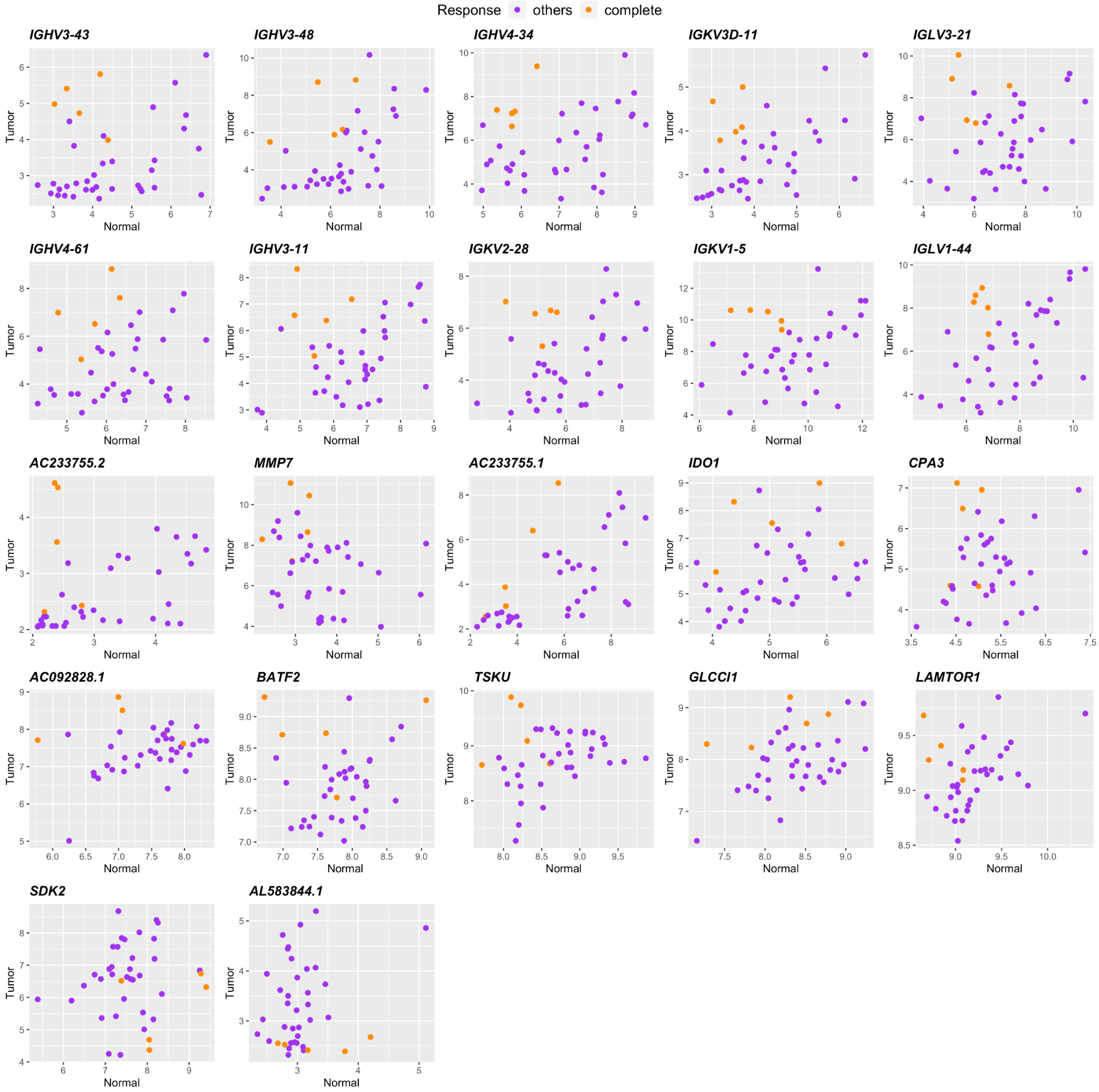


**Supplementary Figure 1. *rlog* values of the differentially expressed genes in tumor samples of complete responders, compared to their corresponding matched normal samples.** We see that for many of these genes, complete responders (orange) cluster at the upper-left quadrant of the plots distinct from other responders (purple) except for SDK2, and AL583844.1 which have negative $\log_2$ fold changes.

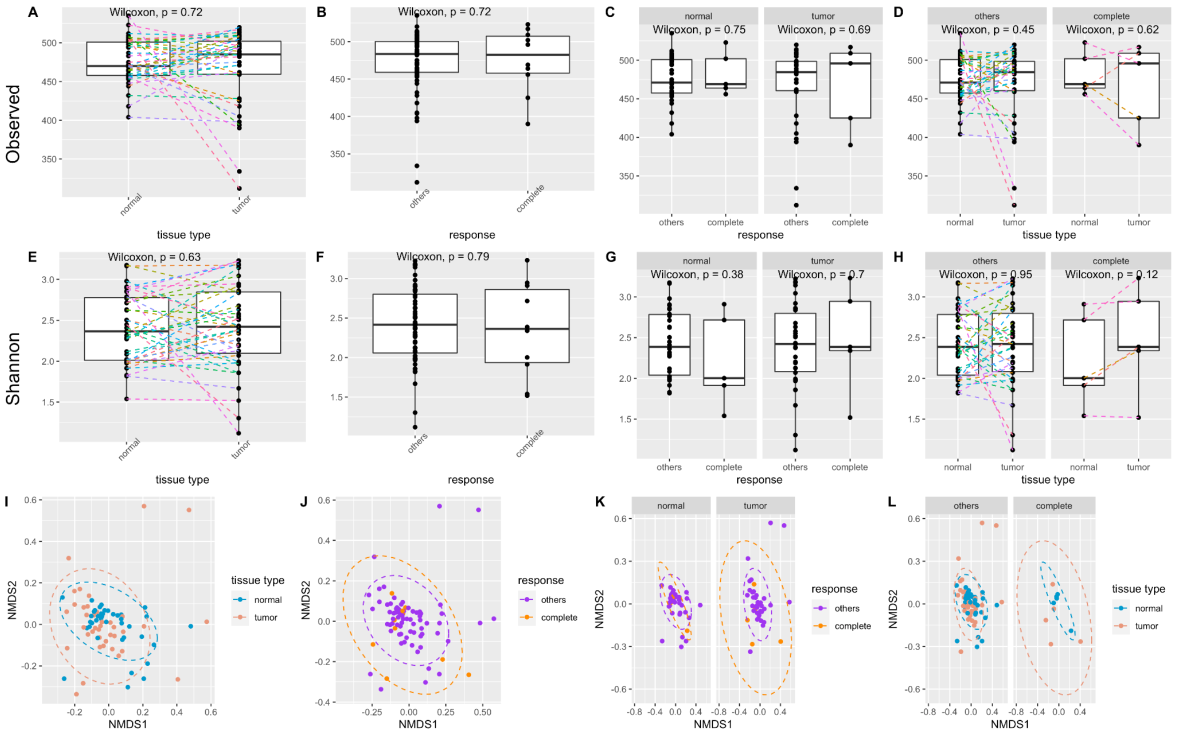


**Supplementary Figure 2. Diversity analyses on different grouping combinations of tumor, normal, and response groups in rectal cancer. A-D:** Observed Alpha Diversity. **A.** between normal and tumor, **B.** between response groups, **C.** between response groups within tissue type, **D.** between tissue type within response groups. **E-H**. Shannon Alpha Diversity. **E.** between normal and tumor, **F.** between response groups, **G.** between response groups within tissue type, **H.** between tissue type within response groups. **I-L**. NMDS ordination of Bray-Curtis distances between samples, grouped by: **I.** between normal and tumors, **J.** between response groups, **K.** between response groups within tissue types, **L.** between tissue type within response groups.

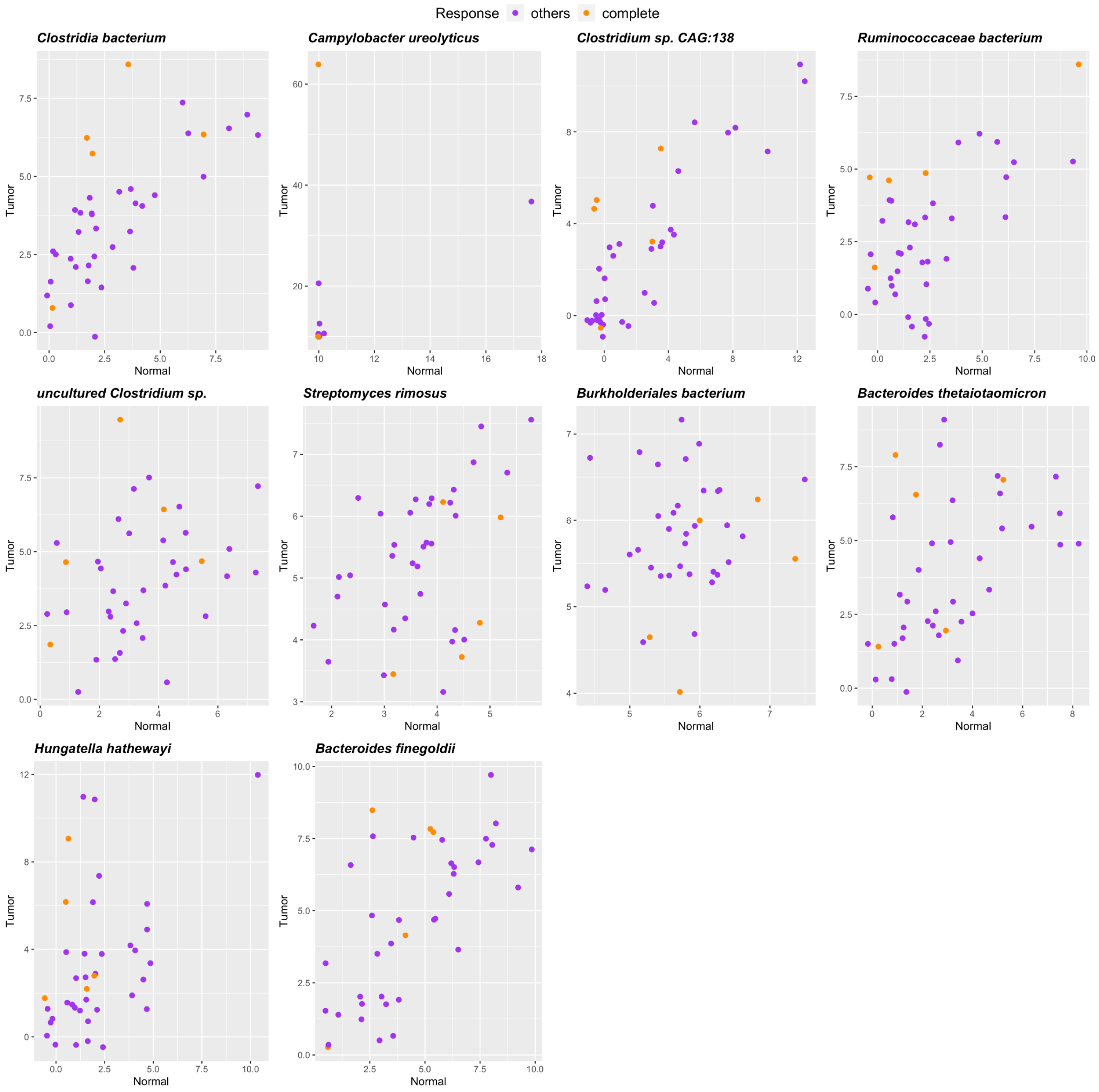


**Supplementary Figure 3. Differentially abundant bacteria in tumor samples compared to matched normal tissue, specific to complete responders**. Scatterplot between rlog values of the bacterial species in tumor samples and their corresponding matched normal samples.
